# Human mobility patterns to inform sampling sites for early pathogen detection and routes of spread: a network modeling and validation study

**DOI:** 10.1101/2024.01.12.24301207

**Authors:** Andrêza L. Alencar, Maria Célia L. S. Cunha, Juliane F. Oliveira, Adriano O. Vasconcelos, Gerson G. Cunha, Ray B. Miranda, Fábio M. H. S. Filho, Corbiniano Silva, Ricardo Khouri, Thiago Cerqueira-Silva, Luiz Landau, Manoel Barral-Netto, Pablo Ivan P. Ramos

**Affiliations:** Department of Computer Science, Federal Rural University of Pernambuco; Recife, 52171-900, Brazil; Luiz Coimbra Institute of Graduate and Engineering Research (COPPE), Federal University of Rio de Janeiro (UFRJ); Rio de Janeiro, 21941-859, Brazil; Center for Data and Knowledge Integration for Health (CIDACS), Gonçalo Moniz Institute, Oswaldo Cruz Foundation (FIOCRUZ); Salvador, 41745-715, Brazil; Rondônia Oswaldo Cruz Foundation, Oswaldo Cruz Foundation (FIOCRUZ); Porto Velho, 76812-245, Brazil; Medicine and Precision Public Health Laboratory (MeSP2), Gonçalo Moniz Institute, Oswaldo Cruz Foundation (FIOCRUZ); Salvador, 40296-710, Brazil

## Abstract

**Background:** Detecting and foreseeing pathogen dispersion is crucial in preventing widespread disease transmission. Human mobility is a critical issue in human transmission of infectious agents. Through a mobility data-driven approach, we determined municipalities in Brazil that could make up an advanced sentinel network, allowing for early detection of circulating pathogens and their associated transmission routes.

**Methods:** We compiled a comprehensive dataset on intercity mobility spanning air, road, and waterway transport, and constructed a graph-based representation of Brazil’s mobility network. The Ford-Fulkerson algorithm, coupled with centrality measures, were employed to rank cities according to their suitability as sentinel hubs.

**Findings:** Our results disentangle the complex transportation network of Brazil, with flights alone transporting 79·9 million (CI 58·3 to 10·1 million) passengers annually during 2017-22, seasonal peaks occurring in late spring and summer, and roadways with a maximum capacity of 78·3 million passengers weekly. We ranked the 5,570 Brazilian cities to offer flexibility in prioritizing locations for early pathogen detection through clinical sample collection. Our findings are validated by epidemiological and genetic data independently collected during the SARS-CoV-2 pandemic period. The mobility-based spread model defined here was able to recapitulate the actual dissemination patterns observed during the pandemic. By providing essential clues for effective pathogen surveillance, our results have the potential to inform public health policy and improve future pandemic response efforts.

**Interpretation:** Our results unlock the potential of designing country-wide clinical sample collection networks using data-informed approaches, an innovative practice that can improve current surveillance systems.

**Funding:** Rockefeller Foundation grant 2023-PPI-007 awarded to MB-N.

**Research in context:** *Evidence before this study:* We searched PubMed on Jun 1, 2023, without language or date restrictions, for the following query: (“mobility network*” OR “transport* network*” OR “sentinel network*” OR “surveillance network*”) AND “model*” AND “surveillance”. The 469 search results were systematically evaluated, and we identified seven original research studies that applied modeling-based approaches to inform the placement, design, or layout of surveillance/sentinel networks. Of these seven studies, four aimed at optimizing the layout of networks for the monitoring of influenza-like illnesses (ILI), while the others aimed at detecting problems arising from the use of medicines based on pharmacy surveillance; detecting the reporting of common acute conditions through a sentinel network of general practitioners; and optimizing the surveillance strategy for plant pests (*S. noctilio*). Most studies employed maximum coverage algorithms that aim to maximize the protected population. Only a single study incorporated mobility patterns to inform the planning of site placement. Studies that involved ILI sentinel networks were geographically restricted to two United States states (Iowa and Texas), and only one study performed a comprehensive whole of United States modeling.

*Added value of this study:* Despite the urgent need to improve the capacity and timeliness of clinical sample collection for public health surveillance, very few studies have tackled the design problem for optimal placement of these sampling sites, and even fewer have used large-scale mobility data to inform these design choices in an epidemiologically-relevant way. Our work contributes to this challenge by leveraging airline/roadway/fluvial mobility data for Brazil that, converted into a graph-based representation and using network metrics, allowed us to pinpoint an optimal layout strategy that could improve the current flu surveillance network of this country. Using data collected during the COVID-19 pandemic, we validated the transmission routes and pathways of SARS-CoV-2 spread, confirming that the mobility data-informed spread scenarios recapitulated the actual dissemination of the virus.

*Implications of all the available evidence:* Mobility data, coupled with network-centered approaches, can complement the identification of strategic locations for early pathogen detection and spread routes.

## Background

Detecting emerging pathogens with pandemic potential represents a significant challenge for health authorities and the scientific community, requiring a deep understanding of various factors, including ecological, evolutionary, climatic, and human behavior^1–6^. In particular, pinpointing specific locations suitable for early pathogen detection and functioning as critical hubs for spread is both critical and complex, and should be a critical feature in next-generation surveillance systems^7^.

Increased human mobility has significantly boosted the risk of new pandemics^6,8–10^. Today’s highly connected world is evidenced by the extensive global transportation network that summed over 38·9 million flights worldwide in 2019^11^. Additionally, there were around 1·39 million interstate bus trips in 2019^12^. Several studies have highlighted the impact of human movement on the spread of infectious diseases, now facilitated by the wealth of available data^8–10^.

Established strategies for anticipating the emergence of a new pathogen involve monitoring clinical and epidemiological data while prioritizing sampling in areas with a high risk of spillover. However, persistent gaps exist in determining optimal study designs, data collection and sharing methods, and sampling intensity to reliably and timely detect signs of pathogen emergence^6,13–15^.

Many countries have created sentinel networks for targeted disease surveillance, with great focus on influenza following the influenza A(H1N1) pandemic of 2009. In Brazil, a surveillance system for flu syndrome relying upon sentinel units was launched nationwide in 2000. Mandatory notification of severe acute respiratory syndrome for hospitalized and deceased cases was implemented in response to the 2009 pandemic. However, akin to many countries, significant challenges limit the representativity of this strategy, including inadequate geographic coverage of sentinel units and *ad hoc* positioning based on existing infrastructure, budget, or mere convenience^16,17^.

This study aims to address the challenges of detecting and tracking the spread of infectious diseases at initial stages in a country with continental proportions. This objective is realized by leveraging mobility data and computational algorithms to identify optimal geographic placement of sampling sites—essentially creating a sentinel network with a data-driven and epidemiologically relevant design. By doing so, pathogens known to constitute a threat to public health can be detected early on, and preemptive actions can be taken before widespread transmission occurs. The integration of mobility data into pandemic surveillance offers a fresh perspective on early warning systems, effectively incorporating epidemiological intelligence into the design and planning process.

## Results

### Spatio-temporal analysis of domestic air transportation

We started by collecting and curating a large-scale dataset for Brazil’s transportation network. From 2017 to 2022, 79·9 million passengers flew annually, representing an average of 39·3% of Brazil’s population. Seasonal peaks in air traffic data occur in late spring and summer. During this period, October counted an average of 7·14 million passengers (8·9% of yearly average flights), 7·24 million passengers in November (9·1%), and 7.96 million in December (10·0%), with January reaching a maximum of 8·33 million passengers (10·4%). A non-seasonal peak of around 6·94 million passengers is observed in July, representing 8·7% of the yearly passenger flow. However, strict mobility measures impacted these counts in 2020 and 2021. In these years, 48·7 and 31·1 million passengers flew, respectively, declines that impacted all 27 federative units of the country.

Understanding the distribution and dynamics of air transportation infrastructure is crucial for evaluating the potential impact on the spread of infectious diseases, and municipalities with operational airports serve as key nodes in the nationwide transportation network. From 2017 to 2023, Brazil averaged 159 such municipalities (out of 5570), with a clear upward trend in the number of municipalities with active flights despite the pandemic-induced decrease in passenger numbers. Most states had fewer than five municipalities with active airports (**Supplementary Figure 1**), with notable exceptions including Pará, the state holding the largest number of municipalities with active airports in 2023 (17 airports).

The Southeastern region of Brazil is a major hub for air travel, hosting an average of 2·4 million incoming passengers (51·3% of all arrivals) and outbound travelers (52·2% of departing passengers) during the examined period. The South follows with 916833 (19·4%)/904268 (19·1%) incoming/departing passengers, respectively. Subsequently, the Central-West registered 681116 (14·4%)/668255 (14·1%) incoming/departing passengers, the Northeast region had 483685 (10·2%)/473309 (10·0%) incoming/departing passengers, and the North region witnessed 227430 (4·8%)/222985 (4·7%) incoming/departing passengers (see **Supplementary Figure 2** and **Supplementary Figure 3**).

Notably, the North and Northeast regions have limited direct connections with the Southern region of Brazil and with each other. Air mobility between these regions may be facilitated by the Southeast and Central-West regions (**Supplementary Figure 3**). Conversely, the Southern region is primarily connected to the Southeast region, with a more fragile connection to the Central-West region. **Figure 1** visualizes the average passenger mobility in air transport, highlighting the influence of states within these regions on the air network.

**Figure 1.**
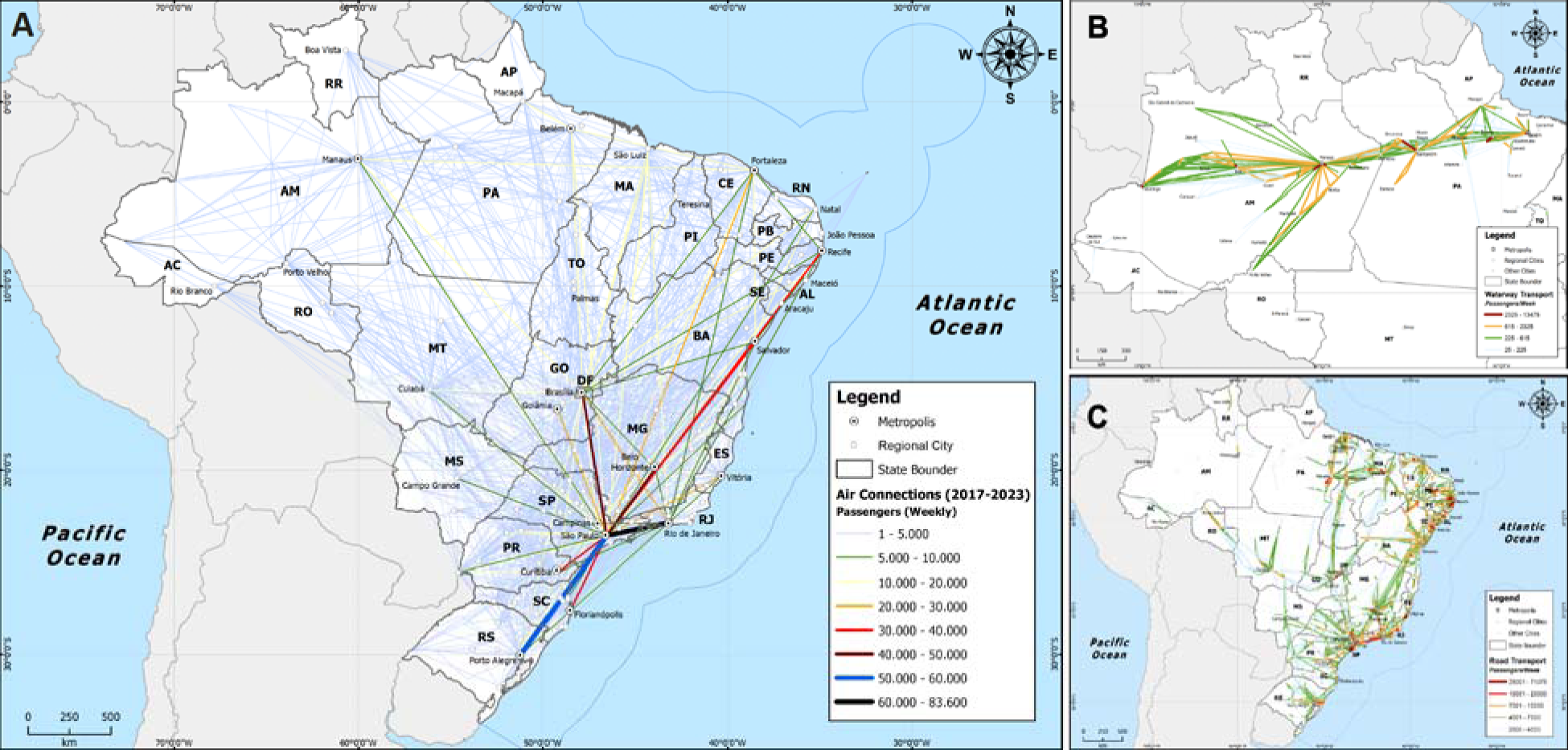
Brazil’s mobility patterns – airway, road, and waterways. A) Average passenger volume in the air mobility network from 2017 to 2023. The average frequency of transport by waterways and roads are shown in panels B and C, respectively. Two-letter state abbreviations are as follows: AC, Acre; AL, Alagoas; AP, Amapá; AM, Amazonas; BA, Bahia; CE, Ceará; DF, Distrito Federal; ES, Espírito Santo; GO, Goiás; MA, Maranhão; MT, Mato Grosso; MS, Mato Grosso do Sul; MG, Minas Gerais; PA, Pará; PB, Paraíba; PR, Paraná; PE, Pernambuco; PI, Piauí; RJ, Rio de Janeiro; RN, Rio Grande do Norte; RS, Rio Grande do Sul; RO, Rondônia; RR, Roraima; SC, Santa Catarina; SP, São Paulo; SE, Sergipe; TO, Tocantins.

### Road and fluvial intercity mobility landscape

We complemented the airline mobility data, which offers monthly totals of passengers flown, with an additional mobility layer that enables the estimation of road/fluvial flows between cities. According to the available data, the country exhibits a weekly mobility capacity of around 78·6 million passengers using these transportation modes, representing 38·7% of the Brazilian population.

Water transport constitutes only 258581 (0.3%) of passenger mobility capacity and is more common in the Northern region of Brazil. In the Amazon state, a substantial 71·4% of mobility capacity relies on river transportation, surpassing road transport. Moreover, this mode of transport forms robust connections, with 99·5% of the link between the states of Amazonas and Pará and 41·3% of the connection between Amazonas and Rondônia facilitated by river transport. Notably, there is a significant water transport connection between the states of Alagoas and Sergipe in Northeastern Brazil, accounting for 12·5% of connectivity. Fluvial transport is also a minority presence in other areas of Brazil, with connections amounting to less than 5%. A representation of passengers’ mobility capacity for both roads and waterways in Brazil is shown in **Figure 1**.

The regional influence on the road and fluvial network also differs from that of air transport. The Northeast region holds the largest share of mobility capacity for passengers at 40·2%, with the Southeast at 28·9%, South at 16·9%, Central-West at 7·6%, and North at 6·1% (**Supplementary Figure 3**).

Similar to the air transportation network, the North and Northeast regions have limited direct connections with the Southern part of Brazil but have more significant connections among themselves (**Supplementary Figure 3**). The Southern region has a more distributed connection with the Southeast and Central-West regions, and the Central-West with the North and Northeast regions compared to the air mobility network (**Supplementary Figure 3**).

### Estimating pathogen pathways and sentinel hubs for early pathogen detection

The previously identified mobility patterns allow for the estimation of routes for the spread of infectious diseases by leveraging network metrics such as the betweenness index (BI) (**Supplementary Text 2**). In a nationwide ranking based on the BI, we identified 1391 cities with BI scores equal to or above the BI upper quartile. Using the Ford-Fulkerson algorithm, we derived 7746479 paths initiating from a node in each Brazilian city and concluding at each distinct city with a high BI score. This approach facilitated categorizing and ranking the primary routes for pathogen transmission, revealing strategic cities for close monitoring that could function as sentinel sampling sites in each state (see Data Availability section).

#### Disentangling transmission routes from key Brazilian cities

We next simulated the emergence of a pathogen initially seeded in four key cities: Manaus (the most populated city in the Amazon region with a population of 2·06 million), Recife (a tourist city in the Northeast with 1·48 million people), Rio de Janeiro and São Paulo, the two largest cities in the country with populations of 6·75 million and 12·3 million, respectively, jointly forming the Megalopolis of the Brazilian Southeast. **Figure 2A** shows the first-step cities in this analysis. Among all paths starting in Manaus and leading to the 1391 cities according to the BI score, 53·6% have São Paulo (city of São Paulo and neighboring municipality Campinas) as their initial destination. This indicates that São Paulo serves as the central primary transmission hub following an emergency in Manaus. Brasília in the Federal District (21·9%), cities in the state of Pará (10·2%), Rondônia (7·8%), Roraima (4·8%), Ceará (0·8%), and Pernambuco (0·5%) are the other most likely states to be affected by a spread emerging in Manaus.

**Figure 2.**
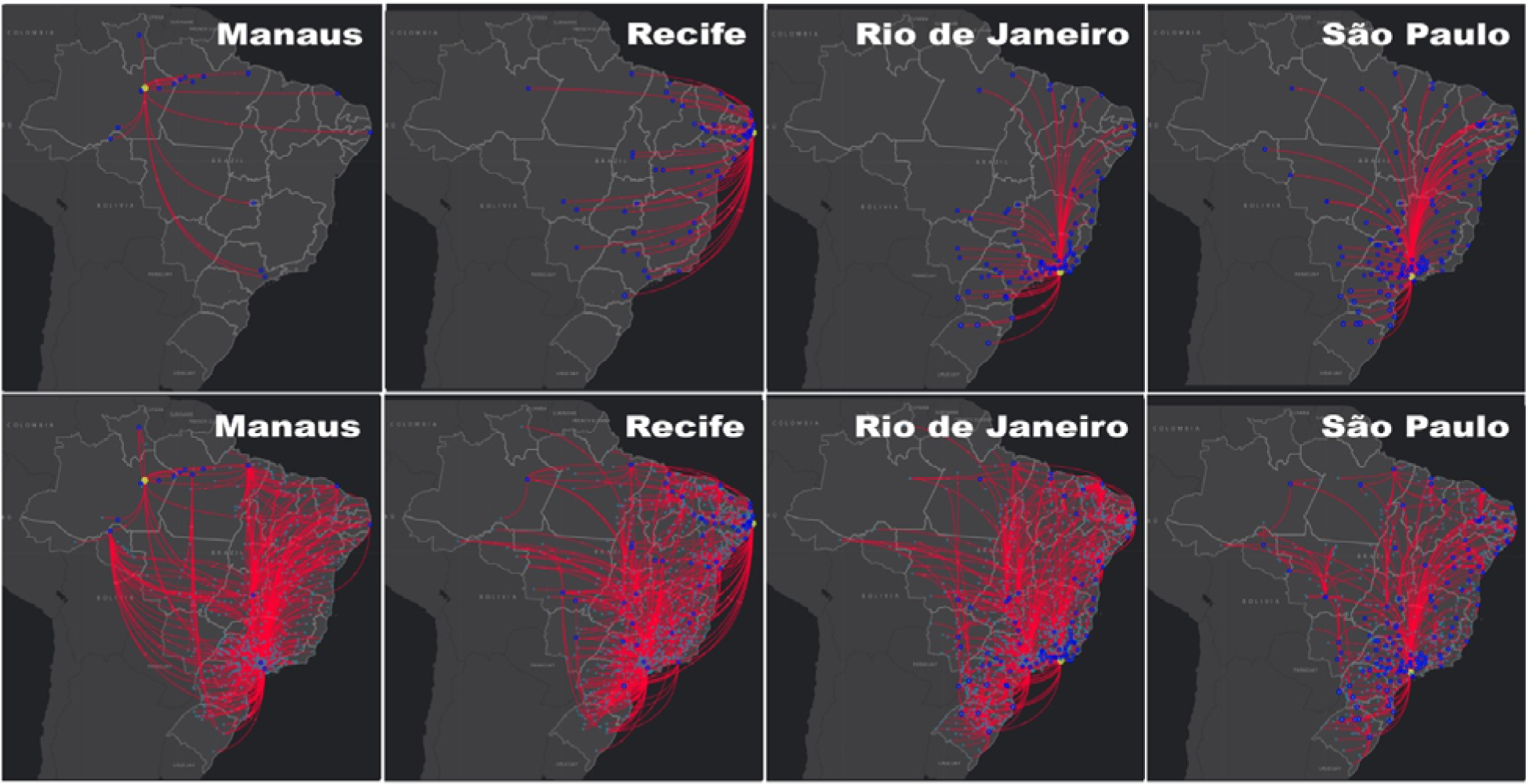
The spread of a pathogen first detected in Manaus/AM, Recife/PE, Rio de Janeiro/RJ, and São Paulo/SP. The maps in the first row depict the initial propagation range for each path originating in the selected city (first-step cities). In the second row, the extension of these paths for second-step cities are shown. In both panels, yellow dots denote the origin, dark blue dots represent first-step cities, and light blue dots represent second-step cities.

Recife and Rio de Janeiro exhibit a wider spread potential to other states, and São Paulo plays a crucial role in the subsequent spread from both Rio de Janeiro and Recife, representing 28·7% and 39·6%, respectively, of the first-step cities in the state of São Paulo. As shown in **Figure 2B**, further spread to the second step of the paths would already lead to extensive transmission across the country.

#### Ranking early warning detection hubs and state gateway cities

Analyzing all pathways starting from cities in each state enables the creation of a ranking that assesses the likelihood of early pathogen detection capacity of each city. This ranking further facilitates the identification of cities according to their significance as mobility gateways to other states. Mobility gateways are defined as cities for which the first step in the most likely paths originating from them leads to cities in other states. Data for the states of Acre, Amazonas, and Rio de Janeiro are presented in **Supplementary Table 1**.

According to the BI score, pre-selected cities in Acre are Rio Branco (the state capital), Cruzeiro do Sul, and Sena Madureira. In the event of a pathogen emergency in a city in Acre that goes undetected, Rio Branco and Cruzeiro do Sul emerge as the most probable cities for pathogen detection using the sentinel approach. Approximately 33·5% and 23·2% of paths starting in Acre use these cities as the first step, respectively. **Figure 3** provides a visual representation of the pathways from cities in Acre to the 1391 selected semi-hubs across the country. Moreover, the list of gateway cities features Rio Branco and Sena Madureira as the key priority points for mobility to other cities outside of Acre.

**Figure 3.**
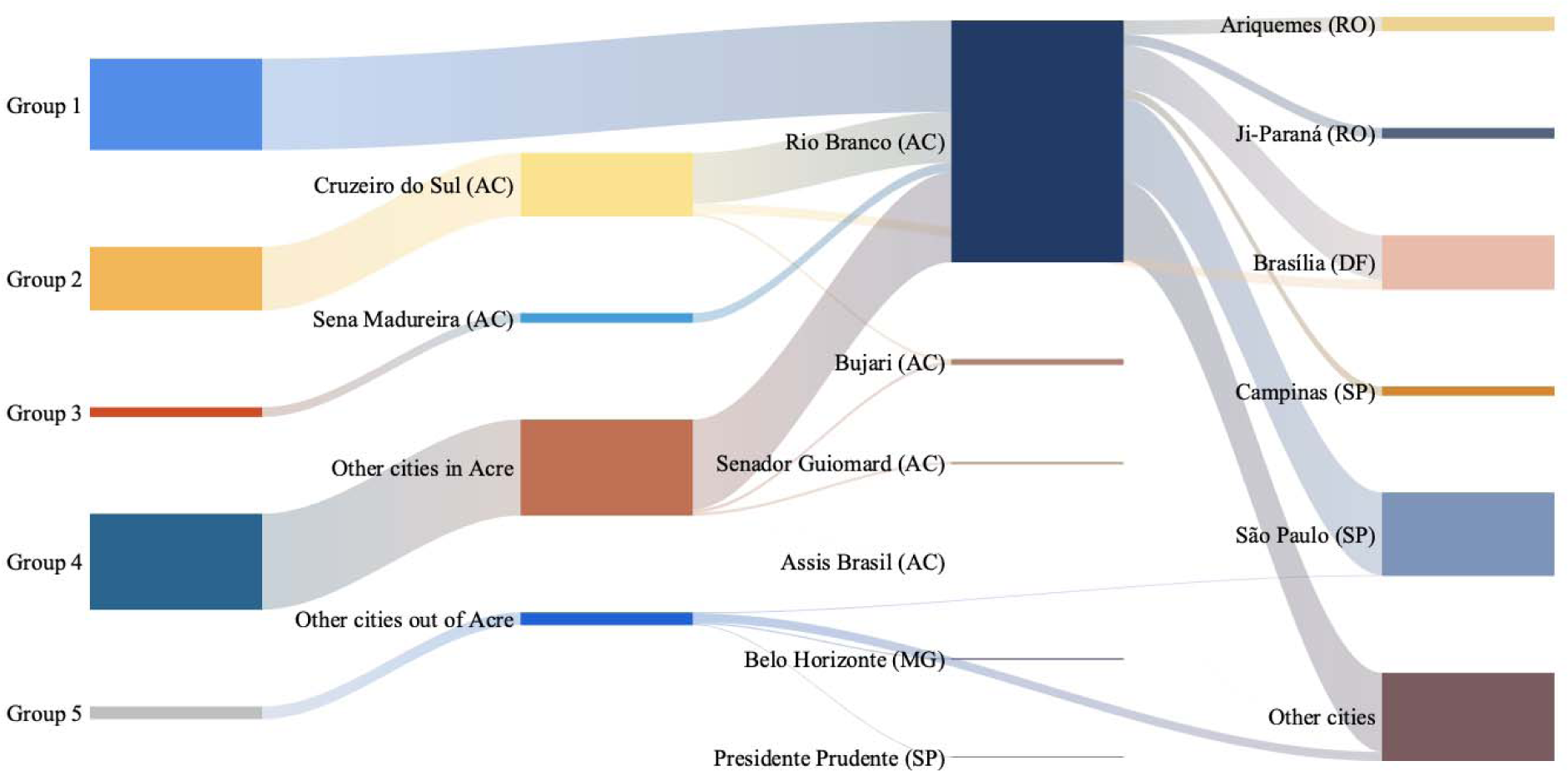
Transmission routes for cities in the state of Acre, North Brazil. Groups 1, 2 and 3 represent cities in Acre, with Rio Branco, Cruzeiro do Sul and Sena Madureira as their respective first-step cities. Group 4 comprises cities with the first step being a non-pre-selected city according to the BI in Acre. Group 5 includes cities with their first step being cities with high BI scores in other states.

Similar to Acre, the priority ranking for early detection cities in the state of Amazonas could not solely be determined by the BI score selection. In this region, the capital city Manaus, followed by Tefé, emerge as the most likely candidates for early pathogen detection. These cities account for 37·7% and 8·6%, respectively, of paths leading to them as the initial step.

Amazonas has 66 cities, with 16 serving as gateways for transmission to other states. These gateway cities can be sorted into two categories according to their BI: The first group, comprising gateway cities with high BI exhibit a broader reach, allowing passenger flows toward more distant states such as São Paulo, the Federal District, Ceará, Pernambuco, as well a neighboring states like Rondônia, Roraima, and Pará. In contrast, gateway cities with low BI scores (falling in the second group) primarily impact neighboring states, including Rondônia, Acre, Roraima, and Pará.

In the case of Rio de Janeiro state, the data reveals a pattern where pre-selected cities according to the BI dominate as the top five candidates for early pathogen detection. The state capital, Rio de Janeiro city, represents a substantial portion, accounting for 40·4% of paths originating from these cities as the initial step. Following a similar analysis as that conducted for Amazonas, we observe that gateway cities in Rio de Janeiro, that satisfy our pre-selection criterion, contribute to a broader transmission spanning 19 states out of the 27. In contrast, gateway cities in Rio de Janeiro that are not pre-selected tend to have a more localized impact, reaching nearby states such as São Paulo, Espírito Santo, and Minas Gerais, as well as more distant states like Tocantins and Bahia.

### Validation with the initial entry period of SARS-CoV-2 in Brazil (February-April, 2020)

During the early stage of the COVID-19 pandemic in Brazil, the first reported cases emerged in the cities of São Paulo and Rio de Janeiro on February 26, 2020 and March 5, 2020, respectively. In this period, a total of 44 and 13 cases of SARS-CoV-2 infection were reported in each city, respectively. Another 19 states had reported cases, although they did not exceed five cases and were mostly related to imported cases at that point. In **Figure 4**, we show the relationship between the proportion of cases in cities in the first, second, and third steps relative to the total number of reported cases in each state until April 1, 2020.

**Figure 4.**
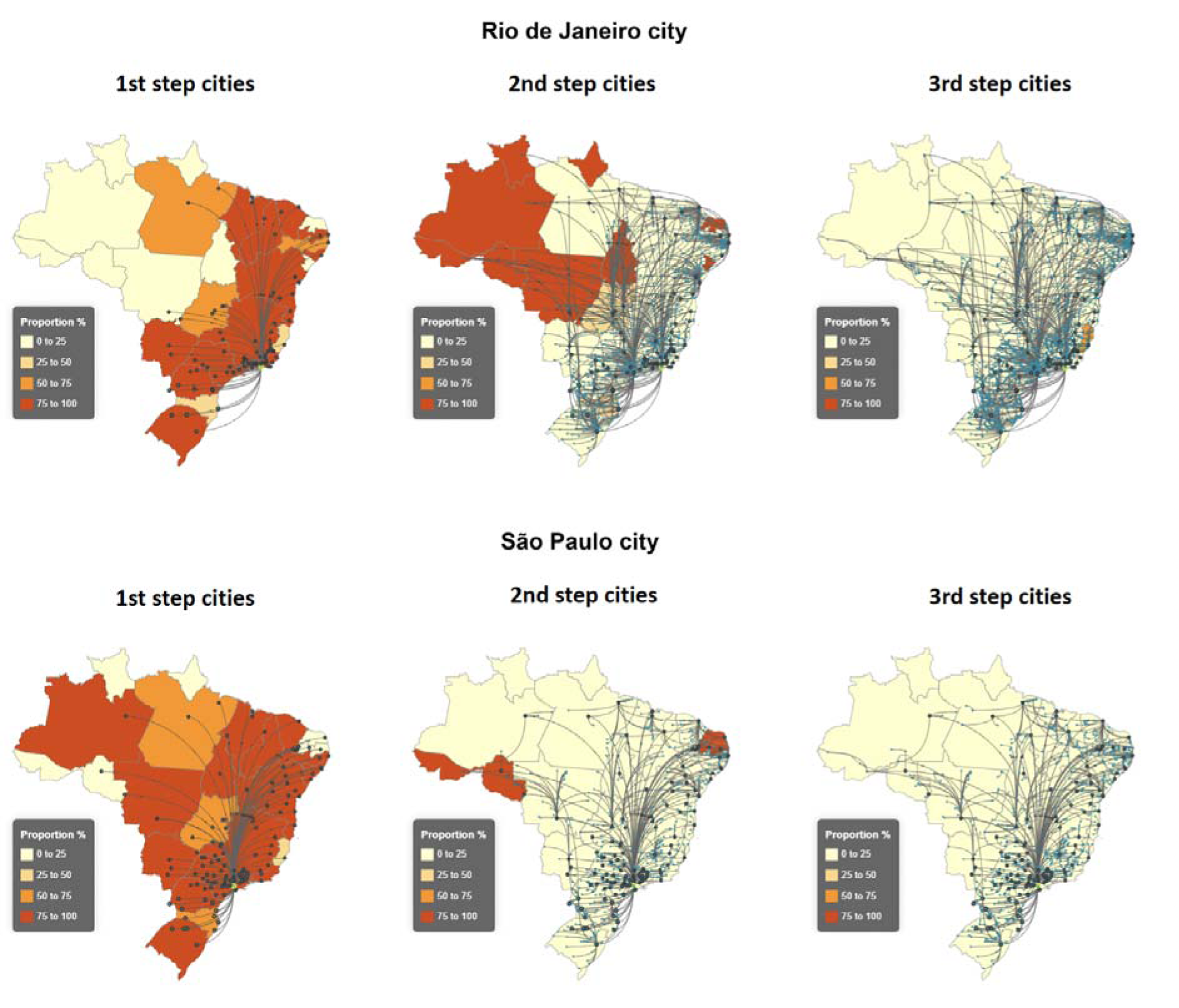
Epidemiological validation of pathogen transmission pathways. Spatial distribution of early-stage (up to April 1, 2020) cities affected by the spread of SARS-CoV-2 originating in the cities of Rio de Janeiro and São Paulo. Brazilian states are colored based on the proportion (%) of COVID-19 cases reported in cities in the first-, second-, and third-steps relative to the total number of reported cases in each state. Lines depict the paths in first step-cities, second-step cities, and third-step cities.

Figure 4 evidences that the 83 first-step cities identified for Rio de Janeiro municipality recapitulate the actual pattern of initial SARS-CoV-2 dissemination during the studied period, with 66·0% of the cases reported in these cities. Most of these first-step cities locate along the Brazilian shoreline (Figure 4). Then, in second-step cities the disease spread towards the North and Mid-West regions, concentrating less connected cities that require additional intermediate connections to be reached.

On the other hand, São Paulo, one of Latin America’s largest cities, presents with a much higher transmission capacity across the country, with first-step cities distributed in more states compared to Rio de Janeiro (Figure 4). Similarly, 83·8% of cases were identified in first step cities. The states of Rondônia and Acre in the North and Paraíba and Rio Grande do Norte in the Northeast are the states with a lower proportion of cases in first-step cities on the paths of São Paulo, in agreement with the strength of connection between these states and the state of São Paulo. Finally, the following steps (2^nd^ and 3^rd^ cities) show a pattern consistent with the observed spatial spread (Figure 4).

### Validation with the initial entry period of the Gamma (P.1) variant in Amazonas (January-March, 2021)

SARS-CoV-2 sequencing data produced during the entry of the Gamma (P.1) variant of concern, which emerged in December, 2020 in the Amazonas state^18^, was also used to validate the sentinel approach. By January 6, 2021, a total of 133 cases were detected in Manaus, with a few sporadic cases detected in other locations (fewer than 13). Table 1 highlights that most cases concentrated in cities at the first step of paths originating from Manaus during the period before February 1, 2021. As the Gamma variant further spread, a larger proportion of cases emerged in cities at the second step. Confirming the data shown for the initial entry of SARS-CoV-2 in Brazil, this additional, real-world result on the spatial distribution of this variant is consistent with the mobility patterns illustrated in Figure 2.

**Table 1.** Case distribution in paths originating from Manaus during the early-stage dissemination of the Gamma (P.1) variant in Brazil.

| Period | Total detected<br>Gamma cases | N. of cases<br>detected in 1<sup>st</sup><br>step cities (%<br>of total<br>detected<br>cases) | N. of cases<br>detected in 2<sup>nd</sup><br>step cities (%<br>of total<br>detected cases) | N. of cases<br>detected in<br>3<sup>rd</sup> step<br>cities (% of<br>total<br>detected<br>cases) | N. of cases<br>detected in<br>other cities (%<br>of total<br>detected cases) |
| --- | --- | --- | --- | --- | --- |
| before<br>January<br>16,2021 | 307 | 224 (73%) | 49 (16%) | 6 (2%) | 28 (9%) |
| before<br>February 1,<br>2021 | 583 | 305 (52%) | 187 (32%) | 26 (4%) | 65 (11%) |
| before March<br>1, 2021 | 2208 | 603 (27%) | 1075 (49%) | 172 (8%) | 358 (16%) |

## Discussion

Human mobility is recognized to have a chief role in the management of outbreaks, evidenced by the travel restrictions enacted during COVID-19^8,19^, and during the 2009 H1N1 pandemic^9^. Here, we unveiled the extensive mobility landscapes of one of the world’s largest countries, Brazil, encompassing air, road, and water transportation routes. Key regions and transport modes significantly contributing to Brazil’s inter-city connectivity were identified. Our analysis delves into the historical dynamics of air transport availability in Brazilian cities, highlighting seasonal variations and examining the impact of the COVID-19 pandemic on air travel. This temporal variation serves as a valuable lens to comprehend the influence of pathogen spillovers and spread, aligning with findings in various studies that have used mobility data to assess the spatial dissemination of diseases^8–10,20^.

In particular, we pursued a ranking strategy leading to a data-informed ordering of cities that could make up a sentinel network for clinical sample collection, enhancing the design of Brazil’s currently used flu syndrome surveillance system. Likewise, other works have attempted to address this problem, notably that of Cheng *et al.,* who formulated this as a multi-objective numeric optimization task, enabling simulation scenarios to be compared and providing a framework for optimal site placement^17^. However, no real-world application of that approach was attempted. Others have used real-world data to adapt local influenza sentinel networks by relying on the concept of maximally covered population, in which the number and location of sentinel sites were chosen to elicit a configuration that maximizes the targeted population^16,21^. However, the catalytic power of human mobility on disease spread was not considered in these works.

On the other hand, the construction of a mobility-driven metapopulation model covering 35 states in the United States has been reported^22^. However, despite common overarching goals, our work presents several methodological differences. While the mobility dataset used by Pei *et al.* focuses on commuting (i.e., journey to work) behavior within and between neighboring counties, we were able to collect a national dataset of transportation patterns covering air/road/fluvial transportation modes. Most importantly, our approach is pathogen-agnostic (i.e., no assumptions around the characteristics of an emerging disease, such as transmission rates, are made, other than that its spread is directly influenced by human flow), while mathematical models such as SIR and extensions thereof^22^ rely on several premises, changes of which can greatly impact the results of these models. Predicting the exact location and time of emergence of a new pathogen is a challenging task. Pathogens can appear unnoticed even in unusual places and remain undetected for an extended period of time before triggering a major outbreak (as with Zika virus, or the different SARS-CoV-2 variants). Our results consider every Brazilian city as a potential site for a spillover and provide guidance on how to respond when a new pathogen emerges.

The primary pathogen transmission routes originating at each Brazilian city were identified by applying the Ford-Fulkerson algorithm to Brazilian large-scale mobility data. Additionally, we disclosed strategic cities for early pathogen detection and recognized gateway cities in each state, offering insights into potential widespread transmission. Our results pointed to distinct patterns in Brazil’s Central-West and Southeastern regions of Brazil, where economically prosperous cities are elemental in facilitating mobility among the Northern, Southern, and Northeastern regions. The well-established infrastructure for human mobility in these regions, marked by airport expansion and modernization efforts, emphasizes the urgency of a strategic early detection plan in this context. Such an approach is particularly crucial to mitigate the spread of diseases in less affluent Northern and Northeastern areas, where early detection strategies must account for the scarcity of infrastructure for pandemic surveillance, laboratory, and sampling in these regions, compared to the South/Southeast.

State capitals should not be the sole focus when determining hub cities for primary sampling. Cities within a state may require an intermediate hub for maximal disease spread, and some cities, with direct and robust connections, may function independently from capitals, influencing mobility to other locations. Finally, ranking gateway cities for further mobility provides valuable insights for planning early detection and containment strategies at the state level. Our results strongly agree with epidemiological and genomic data collected during the entry of the gamma (P.1) variant in North Brazil, evidencing the utility of mobility-based sentinel surveillance placement in this continental-sized country^23–25^.

This study has limitations: The limited availability of more up-to-date mobility data on roads and waterways, estimated only up to 2016. The collected mobility data does not encompass private transport, and inter-city mobility for cities sharing common borders is either underestimated or poorly recorded. One potential strategy to mitigate this limitation is to treat these border cities as a collective unit when planning the deployment of early-alert and pathogen detection systems. Despite these challenges, our work offers valuable insights that can serve as a catalyst for future improvements in mobility data collection methodologies.

Beyond the current scope lies the potential inclusion of variables influencing the selection of cities as hubs for early pathogen detection. Factors such as local health priorities and resources, characteristics favoring the emergence of specific pathogens, the availability of comprehensive health services covering all age groups, significant locations where workers have direct contact with animals, and others could be integrated with our ranking list. This integration could enhance the deployment of sentinel locations for early detection and improve predictions of transmission routes.

## Material and Methods

### Data sources

We collected and curated large-scale mobility data from different administrative sources to enable a comprehensive understanding of inter-city mobility via air, road, and river networks. For validation of the findings, we used epidemiological and genetic circulation data of SARS-CoV-2 within Brazilian municipalities during the Gamma (P.1) period.

#### Mobility data

Inter-city mobility data covering the Brazilian territory was collected from the Brazilian Institute of Geography and Statistics (Instituto Brasileiro de Geografia e Estatística - IBGE) for both road and river networks; air transport data was obtained from the National Civil Aviation Agency (Agência Nacional de Aviação Civil - ANAC)^26,27^.

The IBGE database provides the weekly frequency of transport capacity between city pairs, estimated for 2016. The ANAC database contains extensive historical information on regular and non-regular public air transport services operated by domestic and foreign companies, as well as private flights, in Brazil from 2000 onwards. We used data covering the period of 2017-2023 regarding the number of passengers on domestic flights to allow portraying the air mobility landscape in the country prior to (i.e., 2017-2019), during (i.e., 2020-2021) and after the COVID-19 pandemic period (i.e., 2022-2023).

To align the IBGE and ANAC databases, we used the IBGE’s data collection methodology to convert monthly passenger numbers in ANAC into weekly counts^27,28^. In **Supplementary Text 1** the steps for normalization of these data sources are presented.

### Validation of the analytical framework using SARS-CoV-2 transmission data

The analytical model was validated two-fold using data captured during the COVID-19 pandemic period. First, we collected data from confirmed COVID-19 cases for each Brazilian municipality until May 1, 2020, comprehending the early stage of SARS-CoV-2 entry and spread in the country. The data is openly available and provided by the Brazilian Ministry of Health^29^.

This validation using real-world data during the beginning of the spread aimed to confirm whether the mobility-based spread routes, as determined in our simulations, would recapitulate the actual spread pathways that took place in the initial course of COVID-19 transmission between February 26 and March 12, 2020, before community transmission was declared in the country (March 13, 2020)^20^. Thereafter, transmission rates of the disease were altered due to non-pharmaceutical interventions likely affecting the spread patterns. Then, we evaluated the proportion of cities that reported a case in paths obtained using the Ford-Fulkerson algorithm (described below) until April 1, 2020.

In a second, independent validation approach, we leveraged SARS-CoV-2 genomic data to examine the rises and spreading patterns in COVID-19 cases following the Gamma (P.1) variant emergence in Manaus (North region), which according to existing literature originated in that region^23–25^. We considered three entry/spread periods: 1) from January 6 to January 16, 2021; 2) until February 1, 2021; and 3) until March 1, 2021. Combined, these validation strategies aimed to verify whether the analytical framework employed would recapitulate the geographic spread of a newly-emerged pathogen or a sub-variant for which the affected population would be largely immunologically naïve.

### Construction of the mobility network and estimation of transport flows

The intercity mobility data was used to build a mobility network defined by the graph G = (V, E). Here, *V* = {*v*_l_, *v*_2_, …, *v_n_*} represents a set of n nodes, where each node *v_i_* ∈ *V* corresponds to a city. Additionally, E = {*e*_l_, *e*_2_, …, *e_n_*} represents a set of m edges, where each edge e*_i_* ∈ E holds the frequency of mobility between a pair of cities as an associated weight. The Ford-Fulkerson method computes the maximum flow that can be sent from a designated source node to a designated sink node through a network of interconnected nodes and directed edges^30^. **Supplementary Figure 4** outlines the steps involved in the iterative computation of the Ford-Fulkerson algorithm.

This mobility network, coupled with the Ford-Fulkerson method to estimate flows, were used to infer the most suitable locations for early detection and to track the most likely trajectory of a (newly) emerged pathogen in Brazilian cities. Due to the computational complexity of this task, requiring the evaluation of over 31 million combinations of paths between each pair of cities, we selected a subset of cities based on their network centrality. The Betweenness Index (BI), a global centrality measure, was used as the selection criterion. The cities with BI exceeding the third quartile were sought as destinations for our calculated routes starting in each of the 5570 Brazilian cities. The BI is defined as

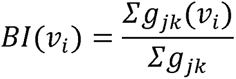

where g represents the number of geodesic paths (shortest paths) between two nodes in the network, and *g_jk_*(*v_i_*) represents the geodesics containing the node *v_i_*, with *i* ≠ *j* ≠ *k* ^31^. We also compared the BI to the proximity index, a secondary centrality metric that is highly correlated with the BI (**Supplementary Text 2**).

### Role of the funding source

The funders had no role in the study design, data collection, analysis, interpretation and writing of this report, nor on the decision to submit the paper for publication.

## Data Availability

All data produced in the present work are contained in the manuscript

https://github.com/andrezaleite/reproducibility_transportation_hubs-early_warning_surveillance_systems.git

https://www.ibge.gov.br

https://www.anac.gov.br

## Acknowledgments

This study is part of the Alert-Early System of Outbreaks with Pandemic Potential (ÆSOP, http://aesop.health), an initiative under development by Brazil’s Fundação Oswaldo Cruz (Fiocruz) and the Federal University of Rio de Janeiro with financial support from the Rockefeller Foundation’s Health Initiative (Grant 2023-PPI-007 awarded to M-BN). The excellent project management and administrative support from Fundação de Apoio à Fiocruz (FIOTEC) is greatly appreciated.

## Author contributions

Conceptualization: ALA, MCLSC, JFO, LL, MB-N, PIPR. Methodology: ALA, MCLSC, JFO, AOV, FMHSF, RK, LL, RBM, PIPR. Investigation: ALA, MCLSC, JFO, AOV, PIPR. Visualization: MCLSC, JFO, AOV, GGC, RBM, CS, PIPR. Funding acquisition: MB-N. Project administration: MB-N, PIPR. Supervision: LL, MB-N, PIPR. Writing – original draft: ALA, JFO, PIPR. Writing – review & editing: all authors.

## Declaration of interests

Authors declare that they have no financial or personal relationships that could be construed as competing interests.

## Data and materials availability

All data needed to evaluate the conclusions in the paper are presented in the paper and/or the accompanying GitHub repository https://github.com/andrezaleite/reproducibility_transportation_hubs-early_warning_surveillance_systems.git. The SARS-CoV-2 genome data used in this study are sourced from GISAID (http://gisaid.org), with accession number gisaid_hcov-19_2022_12_1800. A dashboard enabling direct access to state hubs ranking, gateway cities and paths of transmissions is available and described in the aforementioned GitHub repository, under the *visualization* folder.

## Supplementary Materials

Supplementary Text 1 and 2

Supplementary Figures 1 to 4

Supplementary Table 1

## Notes

### Competing Interest Statement

The authors have declared no competing interest.

### Funding Statement

This study was funded by Rockefeller Foundation grant 2023-PPI-007 awarded to MB-N.

## References

1 Nachega JB, Nsanzimana S, Rawat A, et al. Advancing detection and response capacities for emerging and re-emerging pathogens in Africa. Lancet Infect Dis 2023; 23: e185–9.

2 Edwards AM, Baric RS, Saphire EO, Ulmer JB. Stopping pandemics before they start: Lessons learned from SARS-CoV-2. Science 2022; 375: 1133–9.

3 Taubenberger JK, Kash JC, Morens DM. The 1918 influenza pandemic: 100 years of questions answered and unanswered. Sci Transl Med 2019; 11. DOI:10.1126/scitranslmed.aau5485.

4 Jones KE, Patel NG, Levy MA, et al. Global trends in emerging infectious diseases. Nature 2008; 451: 990–3.

5 Gibb R, Redding DW, Chin KQ, et al. Zoonotic host diversity increases in human-dominated ecosystems. Nature 2020; 584: 398–402.

6 Kraemer MUG, Cummings DAT, Funk S, et al. Reconstruction and prediction of viral disease epidemics. Epidemiol Infect 2018; 147: e34.

7 Ramos PIP, Marcilio I, Bento AI, et al. Combining Digital and Molecular Approaches Using Health and Alternate Data Sources in a Next-Generation Surveillance System for Anticipating Outbreaks of Pandemic Potential. JMIR Public Health Surveill 2024; 10: e47673.

8 Mu X, Yeh AG-O, Zhang X. The interplay of spatial spread of COVID-19 and human mobility in the urban system of China during the Chinese New Year. Environment and Planning B: Urban Analytics and City Science 2021; 48: 1955–71.

9 Bajardi P, Poletto C, Ramasco JJ, Tizzoni M, Colizza V, Vespignani A. Human mobility networks, travel restrictions, and the global spread of 2009 H1N1 pandemic. PLoS ONE 2011; 6: e16591.

10 Peixoto PS, Marcondes D, Peixoto C, Oliva SM. Modeling future spread of infections via mobile geolocation data and population dynamics. An application to COVID-19 in Brazil. PLoS ONE 2020; 15: e0235732.

11 Gullen A, Plungis J. Statista. charleston adv 2013; 15: 43–7.

12 Ministério da Infraestrutura. Ministry of Infrastructure, Brazil. http://portal.infraestrutura.gov.br/ (accessed Dec 20, 2023).

13 Yozwiak NL, Schaffner SF, Sabeti PC. Data sharing: Make outbreak research open access. Nature 2015; 518: 477–9.

14 Quick J, Loman NJ, Duraffour S, et al. Real-time, portable genome sequencing for Ebola surveillance. Nature 2016; 530: 228–32.

15 Grubaugh ND, Ladner JT, Kraemer MUG, et al. Genomic epidemiology reveals multiple introductions of Zika virus into the United States. Nature 2017; 546: 401–5.

16 Polgreen PM, Chen Z, Segre AM, Harris ML, Pentella MA, Rushton G. Optimizing influenza sentinel surveillance at the state level. Am J Epidemiol 2009; 170: 1300–6.

17 Cheng Q, Collender PA, Heaney AK, et al. The DIOS framework for optimizing infectious disease surveillance: Numerical methods for simulation and multi-objective optimization of surveillance network architectures. PLoS Comput Biol 2020; 16: e1008477.

18 Lamarca AP, de Almeida LGP, Francisco R da S, et al. Genomic surveillance of SARS-CoV-2 tracks early interstate transmission of P.1 lineage and diversification within P.2 clade in Brazil. PLoS Negl Trop Dis 2021; 15: e0009835.

19 Chinazzi M, Davis JT, Ajelli M, et al. The effect of travel restrictions on the spread of the 2019 novel coronavirus (COVID-19) outbreak. Science 2020; 368: 395–400.

20 Jorge DCP, Rodrigues MS, Silva MS, et al. Assessing the nationwide impact of COVID-19 mitigation policies on the transmission rate of SARS-CoV-2 in Brazil. Epidemics 2021; 35: 100465.

21 Fairchild G, Polgreen PM, Foster E, Rushton G, Segre AM. How many suffice? A computational framework for sizing sentinel surveillance networks. Int J Health Geogr 2013; 12: 56.

22 Pei S, Teng X, Lewis P, Shaman J. Optimizing respiratory virus surveillance networks using uncertainty propagation. Nat Commun 2021; 12: 222.

23 Naveca FG, Nascimento V, de Souza VC, et al. COVID-19 in Amazonas, Brazil, was driven by the persistence of endemic lineages and P.1 emergence. Nat Med 2021; 27: 1230–8.

24 Giovanetti M, Slavov SN, Fonseca V, et al. Genomic epidemiology of the SARS-CoV-2 epidemic in Brazil. Nat Microbiol 2022; 7: 1490–500.

25 Gräf T, Bello G, Naveca FG, et al. Phylogenetic-based inference reveals distinct transmission dynamics of SARS-CoV-2 lineages Gamma and P.2 in Brazil. iScience 2022; 25: 104156.

26 Brazilian National Civil Aviation Agency (ANAC). https://www.anac.gov.br (accessed Dec 20, 2023).

27 Instituto Brasileiro de Geografia e Estatística. Coordenac□Jão de Geografia,. Ligac□Jões Rodoviárias E Hidroviárias, 2016. Rio de Janeiro: Rio de Janeiro□J: IBGE, Instituto Brasileiro de Geografia e Estatística, 2017.

28 Brazilian National Transport Confederation. https://anuariodotransporte.cnt.org.br/2022/Inicial (accessed Dec 20, 2023).

29 Ministry of Health, Brazil. Informatics Department Open Data SUS System. https://covid.saude.gov.br/ (accessed Dec 20, 2023).

30 Ford LR, Fulkerson DR. Maximal flow through a network. Canad J Math 1956; 8: 399– 404.

31 Nakamura H, Managi S. Airport risk of importation and exportation of the COVID-19 pandemic. Transport Policy 2020; 96: 40–7.

