## Supplementary materials for "Human mobility patterns to inform sampling sites for early pathogen detection and routes of spread: a network modeling and validation study"

**The PDF file includes:**

Supplementary Text 1 and 2

Supplementary Figures 1 to 4

Supplementary Table 1

**Supplementary Text 1**

IBGE methodology

To conduct a study integrating IBGE and ANAC databases, we used the IBGE's data collection methodology to convert monthly passenger numbers in ANAC to weekly counts.

The IBGE methodology relies on data from multiple sources, such as questionnaires at bus terminals, commercial ticket offices, municipal bus stops, city halls, waterway terminals, boat cooperatives, individual boat operators, and direct communication with companies. To ensure comprehensive data coverage, they also evaluated informal and alternative transport modes like vans, station wagons, and minibusses.

To standardize the data and enable meaningful comparisons, the IBGE research group converted the weekly frequency of vehicles into a common unit of measurement. First, it calculated the weekly frequency by summing up the number of weekly departures between each pair of municipalities. For cases with only quarterly or monthly frequencies, they adjusted the sum by multiplying it by 0.5 or 0.25, respectively, to match the weekly frequency (27).

Next, it was assigned a default value of 1 to buses, considering them as the baseline measure. Other vehicle types, such as vans and cars, were adjusted by multiplying their frequencies by 0.25. For waterway vehicles, IBGE research group used multipliers to convert their frequencies to the equivalent of a bus. For instance, as described in (27) flying boats had a frequency multiplier of 0.25, speedboats and catamarans were considered equivalent to buses (value 1), boats were multiplied by 1.5, and ships by 2. This approach allowed us to estimate the number of passengers traveling between cities of 25 people per bus, based on the National Transport Confederation (28).

**Supplementary Text 2**

Betweenness and proximity indices

The proximity index (PI) of a node $v_i$ is calculated as the ratio:

$$PI(v_i) = \frac{n-1}{\Sigma D(v_i, v_j)}$$

where n is the total number of nodes and D is a topological distance that measure the number of steps between the nodes $v_i$ and $v_j$, for $i \neq j$.

The PI index ranges from 0 to 1, with 1 indicating that a city is directly connected to another city in the network. In our study, the PI varies between 0 and 0.56, with an average value of 0.31 (SD = 0.07). The cities with the highest values are predominantly located in the center and southeast of Brazil. São Paulo - SP ranks at the top with an index of 0.56, followed closely by the Brazilian capital, Brasilia - BR (0.49), Campinas - SP (0.49), and Goiania - GO (0.49). In contrast, the cities in the northern region of Brazil are more distant from the rest in our network.

The BI exhibits patterns comparable to the PI results but with notable exceptions, as it emphasizes the role of cities in mediating other connections within the network. BI scores vary among cities from 0 to 0.28, with an average of 0.0004 (SD = 0.0044). São Paulo maintains its position as the city with the highest BI value (0.28), followed now by Belo Horizonte (0.07), Goiania (0.05), and Brazilia (0.04). Unlike the PI, the BI highlights important connections in the north and south of the country.

By selecting cities in a nationwide ranking with PI scores equal to or above the third quartile (≥ 0.36), we identify 1,499 cities eligible as influential by this index. The number of cities per state is presented in Table 1. We can see from the selection that states with over 100 cities are Bahia, Minas Gerais, Paraná, and São Paulo. However, northern states like Amapá, Amazonas, and Roraima do not have any cities listed among the most influential in a nationwide PI ranking selection.

By analyzing the BI in a nationwide ranking, we identify 1,391 cities with BI scores equal to or above the third quartile (>=0.000053). Similar to the Proximity Index (PI), we observe that, in addition to the states of Rio Grande do Sul, Bahia, Minas Gerais, Paraná, and São Paulo have more than 100 cities in the selected ranking. Notably, the BI Index reveals cities in all states, especially in regions with more distant cities (low PI values), such as the northern states (Amapá, Amazonas, Pará, Rondônia, Roraima) and the Northeast (Maranhão, Tocantins, Rio Grande do Norte), Southeast (Espírito Santo), and South (Santa Catarina and Rio Grande do Sul). See Table 1 for more details.

**Table 1:** Number of influential cities per State based on proximity and betweenness indexes scores above the third quantile.

| **State name** | **Number of cities by PI (≥ 0.36).** | **Number of cities by BI (≥ 0.000053).** | **Number of cities that coincides in the selected cities by both indexes** | **Proportion of cities relative to the total number of cities in the state** |
|---|---|---|---|---|
| Acre (AC) | 2 | 3 | 2 | 13.6% (3/22) |
| Alagoas (AL) | 22 | 17 | 14 | 16.7%(17/102) |
| Amapá (AP) | - | 1 | 0 | 6.2% (1/16) |
| Amazonas (AM) | - | 5 | 5 | 8.1%(5/62) |
| Bahia (BA) | 182 | 108 | 103 | 25.9% (108/417) |
| Ceará (CE) | 59 | 54 | 49 | 29.3% (54/184) |
| Espírito Santo (ES) | 15 | 25 | 14 | 32.1% (25/78) |
| Goiás (GO) | 39 | 47 | 37 | 19.1% (47/246) |
| Maranhão (MA) | 25 | 50 | 23 | 23.0% (50/217) |
| Mato Grosso (MT) | 39 | 53 | 37 | 37.6% (53/141) |
| Mato Grosso do Sul (MS) | 26 | 28 | 22 | 35.4% (28/79) |
| Minas Gerais (MG) | 259 | 227 | 184 | 26.6% (227/853) |
| Pará (PA) | 14 | 35 | 14 | 24.3% (35/144) |

| | | | | |
|---|---|---|---|---|
| Paraíba (PB) | 48 | 44 | 37 | 19.7% (44/223) |
| Paraná (PR) | 101 | 104 | 79 | 26.1% (104/399) |
| Pernambuco (PE) | 67 | 60 | 58 | 32.4% (60/185) |
| Piauí (PI) | 68 | 53 | 48 | 23.7% (53/224) |
| Rio de Janeiro (RJ) | 30 | 27 | 22 | 29.3% (27/92) |
| Rio Grande do Norte (RN) | 12 | 17 | 12 | 10.2% (17/167) |
| Rio Grande do Sul (RS) | 45 | 110 | 45 | 22.1% (110/497) |
| Rondônia (RO) | 9 | 13 | 9 | 25.0% (13/52) |
| Roraima (RR) | - | 2 | 0 | 13.3% (2/15) |
| Santa Catarina (SC) | 53 | 77 | 52 | 26.1% (77/295) |
| São Paulo (SP) | 353 | 190 | 186 | 29.5% (190/645) |
| Sergipe (SE) | 17 | 16 | 15 | 21.3% (16/75) |
| Tocantins (TO) | 13 | 24 | 13 | 17.3% (24/139) |
| Distrito Federal (DF) | 1 | 1 | 1 | 100% (1/1) |
| **Total** | **1,499** | **1,391** | **1,076** | **25.0% (1391/5570)** |

Due to the high correlation between the indices and a better representativeness of the influential cities by the BI in all states, the list of selected hubs corresponds to those of the BI index. Therefore, the pre-selected in this subsection are used to employ our FF algorithm.

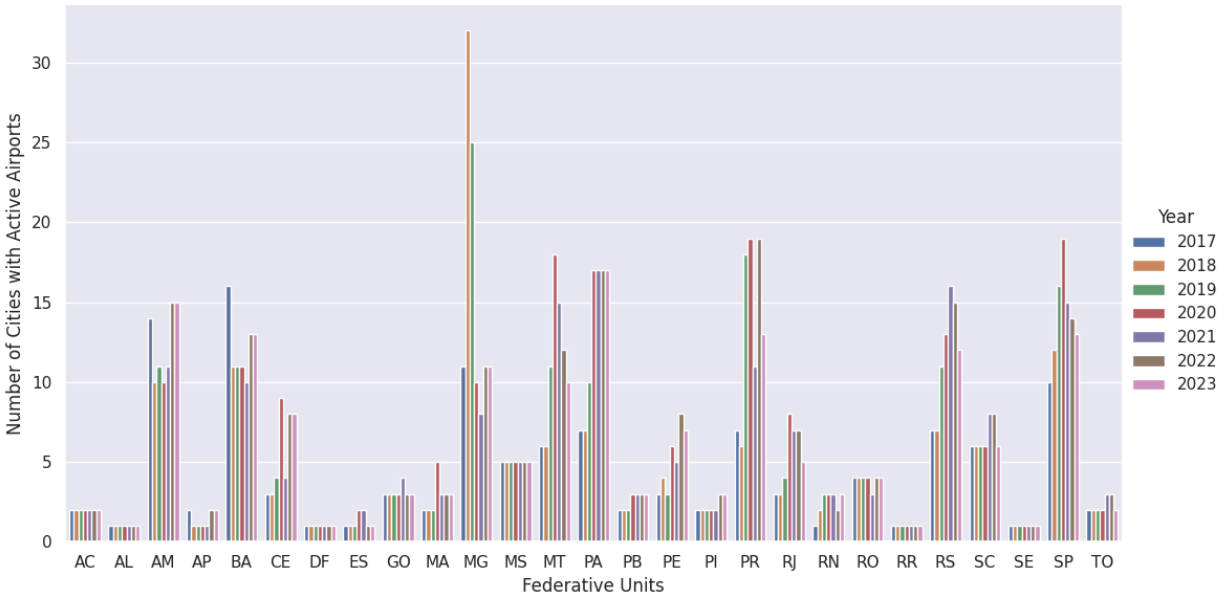


**Fig. S1. Progression of cities with active flights by state in Brazilian states from 2017 to 2023.** Two-letter state abbreviations are as follows: AC, Acre; AL, Alagoas; AP, Amapá; AM, Amazonas; BA, Bahia; CE, Ceará; DF, Distrito Federal; ES, Espírito Santo; GO, Goiás; MA, Maranhão; MT, Mato Grosso; MS, Mato Grosso do Sul; MG, Minas Gerais; PA, Pará; PB, Paraíba; PR, Paraná; PE, Pernambuco; PI, Piauí; RJ, Rio de Janeiro; RN, Rio Grande do Norte; RS, Rio Grande do Sul; RO, Rondônia; RR, Roraima; SC, Santa Catarina; SP, São Paulo; SE, Sergipe; TO, Tocantins.. Create a page break and paste in the figure above the caption.

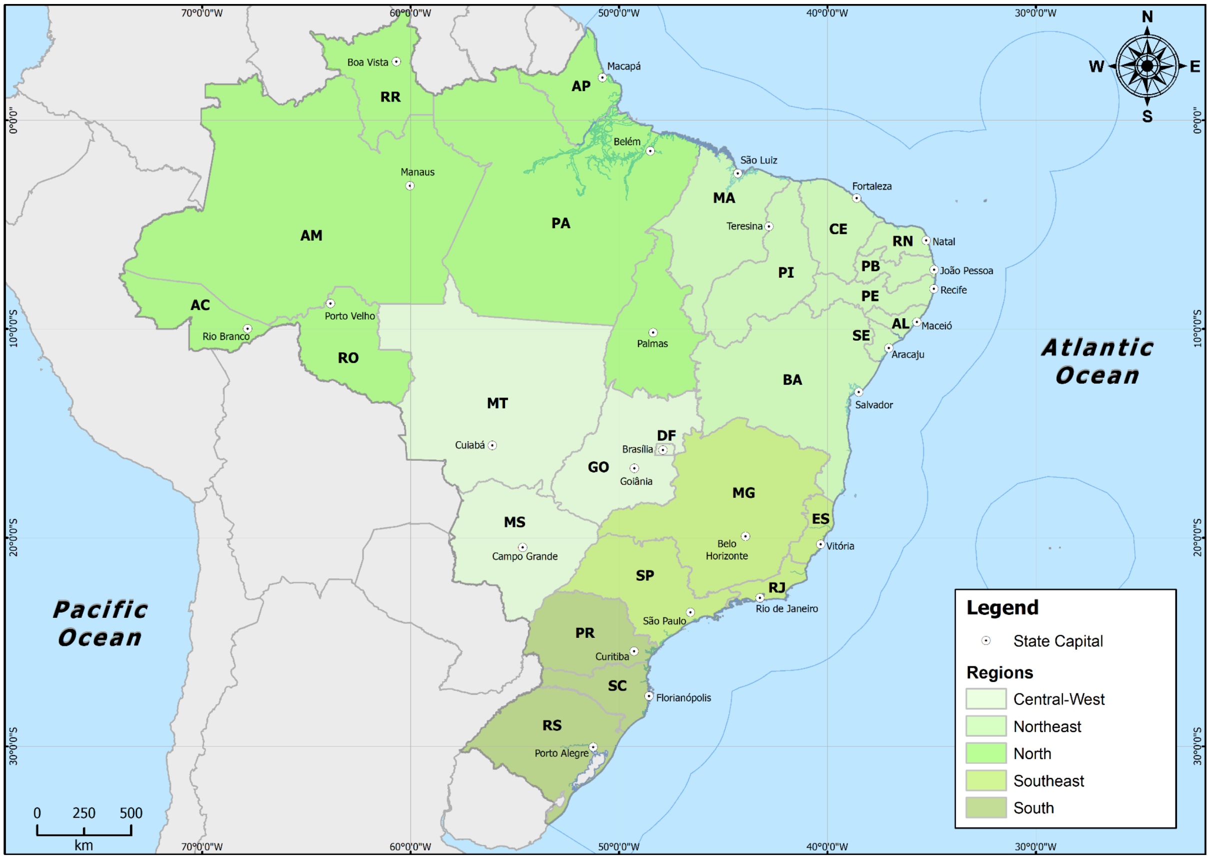


**Fig. S2. Brazilian regions and the states belonging to them.** Two-letter state abbreviations are as follows: AC, Acre; AL, Alagoas; AP, Amapá; AM, Amazonas; BA, Bahia; CE, Ceará; DF, Distrito Federal; ES, Espírito Santo; GO, Goiás; MA, Maranhão; MT, Mato Grosso; MS, Mato Grosso do Sul; MG, Minas Gerais; PA, Pará; PB, Paraíba; PR, Paraná; PE, Pernambuco; PI, Piauí; RJ, Rio de Janeiro; RN, Rio Grande do Norte; RS, Rio Grande do Sul; RO, Rondônia; RR, Roraima; SC, Santa Catarina; SP, São Paulo; SE, Sergipe; TO, Tocantins.

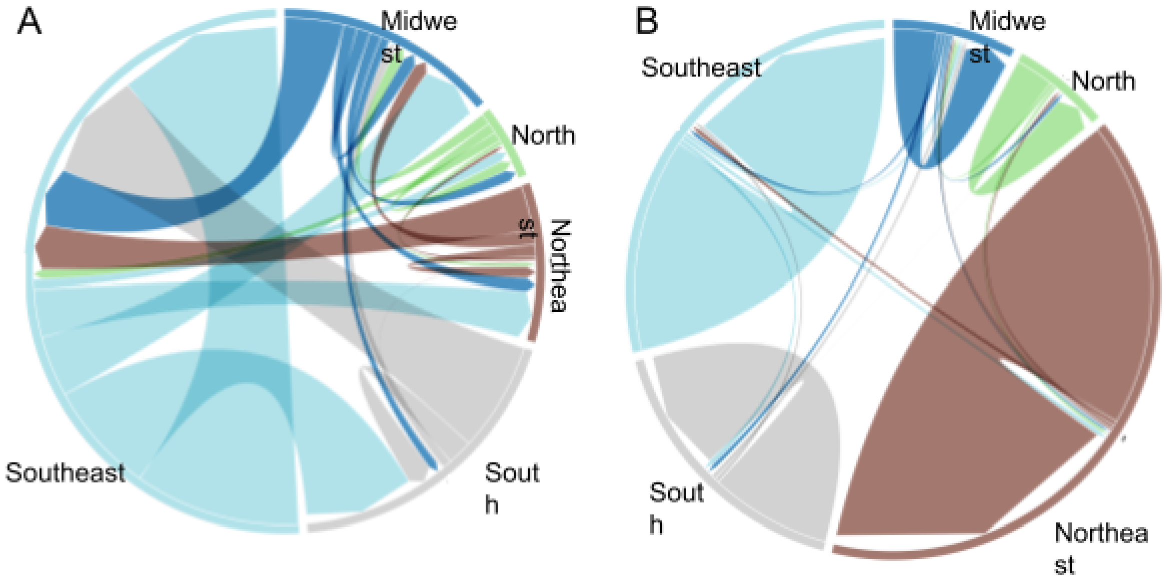


**Fig. S3. Passenger mobility among Brazilian regions**. Inter and extra-region connections are represented by (A) average air mobility and (B) road and waterway.

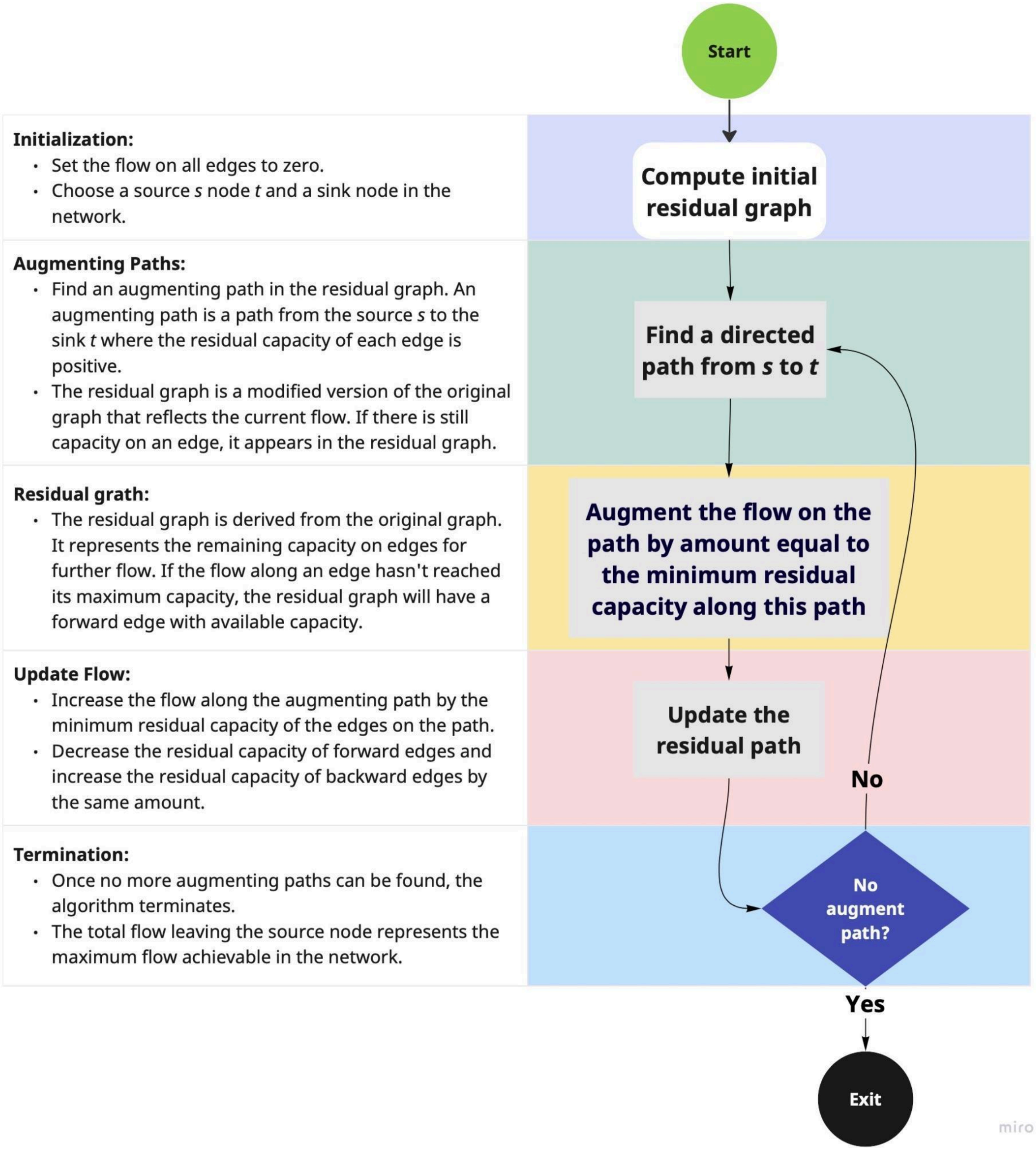


**Fig. S4: Computation iteration of the Ford-Fulkerson algorithm.** Schematic representation of the Ford-Fulkerson algorithm applied to optimize intercity mobility paths for early pathogen detection.

**Table S1: Sentinel hubs according to mobility patterns in Brazil.** Ranking of early warning detection hubs and gateway cities for inter-state mobility in Acre, Amazonas, and Rio de Janeiro.

| State | Ranking of the five cities most likely to be the first step toward early detection in the state | Gateway cities for spread to other states |
|---|---|---|
| Acre | 1º Rio Branco**<br>2º Cruzeiro do Sul**<br>3º Senador Guiomard<br>4º Brasiléia<br>5º Bujari<br>* | 1º Rio Branco**<br>2º Sena Madureira** |
| Amazonas | 1º Manaus**<br>2º Tefé<br>3º Manacapuru<br>4º Itacoatiara**<br>5º Itapiranga<br>* | 1º Boca do Acre<br>2º Apuí<br>3º Guajará<br>4º Ipixuna<br>5º Manaus<br>* |
| Rio de Janeiro | 1º Rio de Janeiro**<br>2º Campos dos Goytacazes**<br>3º Barra Mansa**<br>4º Macaé**<br>5º Três Rios**<br>* | 1º Rio de Janeiro**<br>2º Niterói**<br>3º Sapucaia<br>4º Itatiaia<br>5º Laje do Muriaé**<br>* |
| *Full list available at https://aesop.outerlamce.com/sankey.<br>**Pre-selected cities. | | |
